# Quantifying early COVID-19 outbreak transmission in South Africa and exploring vaccine efficacy scenarios

**DOI:** 10.1101/2020.04.23.20077297

**Authors:** Z. Mukandavire, F. Nyabadza, N. J. Malunguza, D. F. Cuadros, T. Shiri, G. Musuka

**Author notes:** Address for correspondence: Godfrey Musuka, ICAP at Columbia University, Harare, Zimbabwe.

## Abstract

**Background:** COVID-19 has emerged and spread at great speed globally and has presented one of the greatest public health challenges in modern times with no proven cure or vaccine. Africa is still early in this epidemic, therefore the spectrum of disease severity is not yet clear.

**Methods:** We used a mathematical model to fit to the observed cases of COVID-19 in South Africa to estimate the basic reproductive number and critical vaccination coverages to control the disease for different hypothetical vaccine efficacy scenarios. We also estimated the percentage reduction in effective contacts due to the social distancing measures implemented.

**Results:** Early model estimates show that COVID-19 outbreak in South Africa had a basic reproductive number of 2.95 (95% credible interval [CrI] 2.83-3.33). A vaccine with 70% efficacy had the capacity to contain COVID-19 outbreak but at very higher vaccination coverage 94.44% (95% Crl 92.44-99.92%) with a vaccine of 100% efficacy requiring 66.10% (95% Crl 64.72-69.95%) coverage. Social distancing measures put in place have so far reduced the number of social contacts by 80.31% (95% Crl 79.76-80.85%).

**Conclusions:** Findings suggest a highly efficacious vaccine would have been required to contain COVID-19 in South Africa. Therefore, the current social distancing measures to reduce contacts will remain key in controlling the infection in the absence of vaccines and other therapeutics.

## Introduction

The Coronavirus Disease 2019 (COVID-19) originated in Wuhan, China, in December 2019 and has rapidly spread around the world [1]. There is limited understanding of the epidemiology of SARS-CoV-2, the pathogen that causes the disease COVID-19. Currently, the epicentre of the virus is in Europe and New York in the United States of America [2]. In Africa, the virus is just starting to set its foothold, with South Africa now reporting the majority of cases. The first case of the COVID-19 in South Africa was reported on the 5^th^ of March 2020 [3]. Measures to contain the epidemic culminated in the declaration of the state of disaster leading to a national *lockdown* on the 26^th^ of March 2020 with Gauteng, Western Cape, KwaZulu-Natal and the Free State provinces reporting most of the COVID-19 cases. The map in Figure 1 shows the distribution of COVID-19 confirmed cases in South Africa before the government mandated a *lockdown*. Gauteng province appeared to be the “*epicentre*” of COVID-19 in South Africa for a number of reasons. First, the province has the largest population density [4, 5] and the urban population is poor with 20% of its population being food insecure [4]. Second, Gauteng province has two international airports including OR Tambo International Airport handling over 21 million passengers annually [6]. Third, the volume of people that use public transport runs into millions daily creating a fertile ground accelerating the spread of the disease. Finally, the province is the country’s economic hub, and many people (including international visitors) travel in and out of the province daily [4]. With the majority of confirmed cases early in the outbreak having been linked to international travel [7], it is not surprising that the most affected provinces (Gauteng, Western Cape and KwaZulu-Natal) have international airports with direct flights to affected global regions (Figure 1). The cases in the Free State province have mainly been attributed to a cluster transmission resulting from a mega church gathering [8].

**Figure 1:**
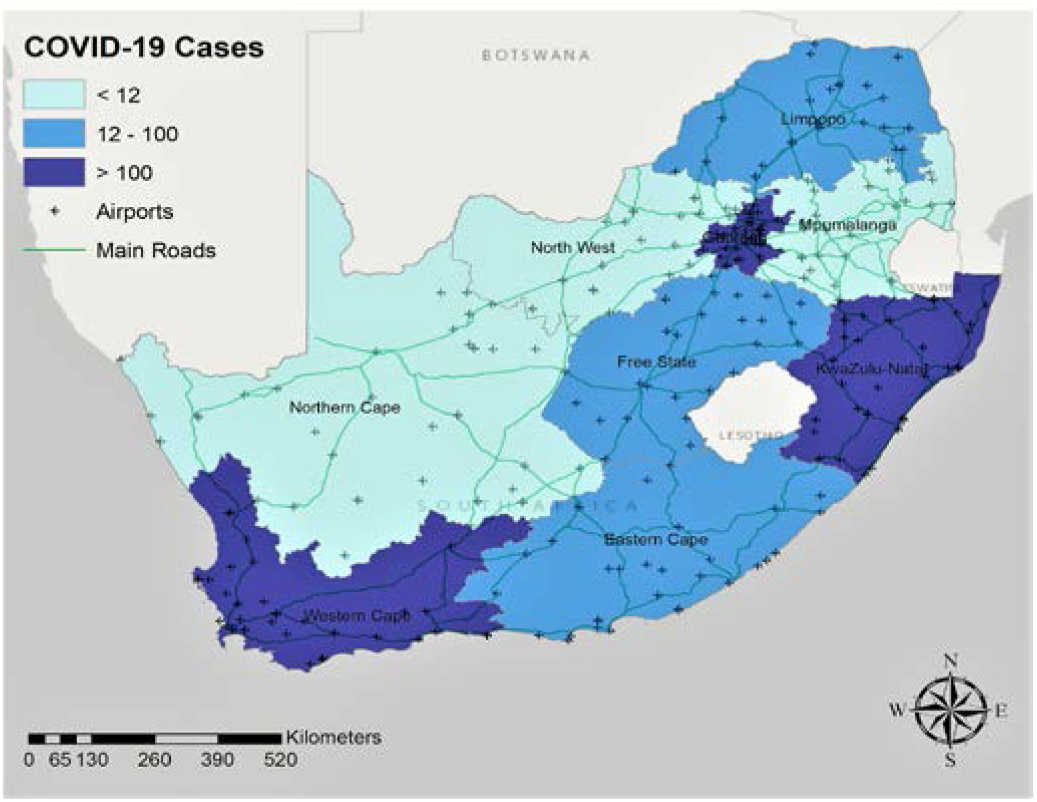
COVID-19 cases distribution in South Africa by 27 March 2020. Map was created using ArcGIS^®^ by ESRI version 10.5 (*http://www.esri.com*).

**Figure 2:**
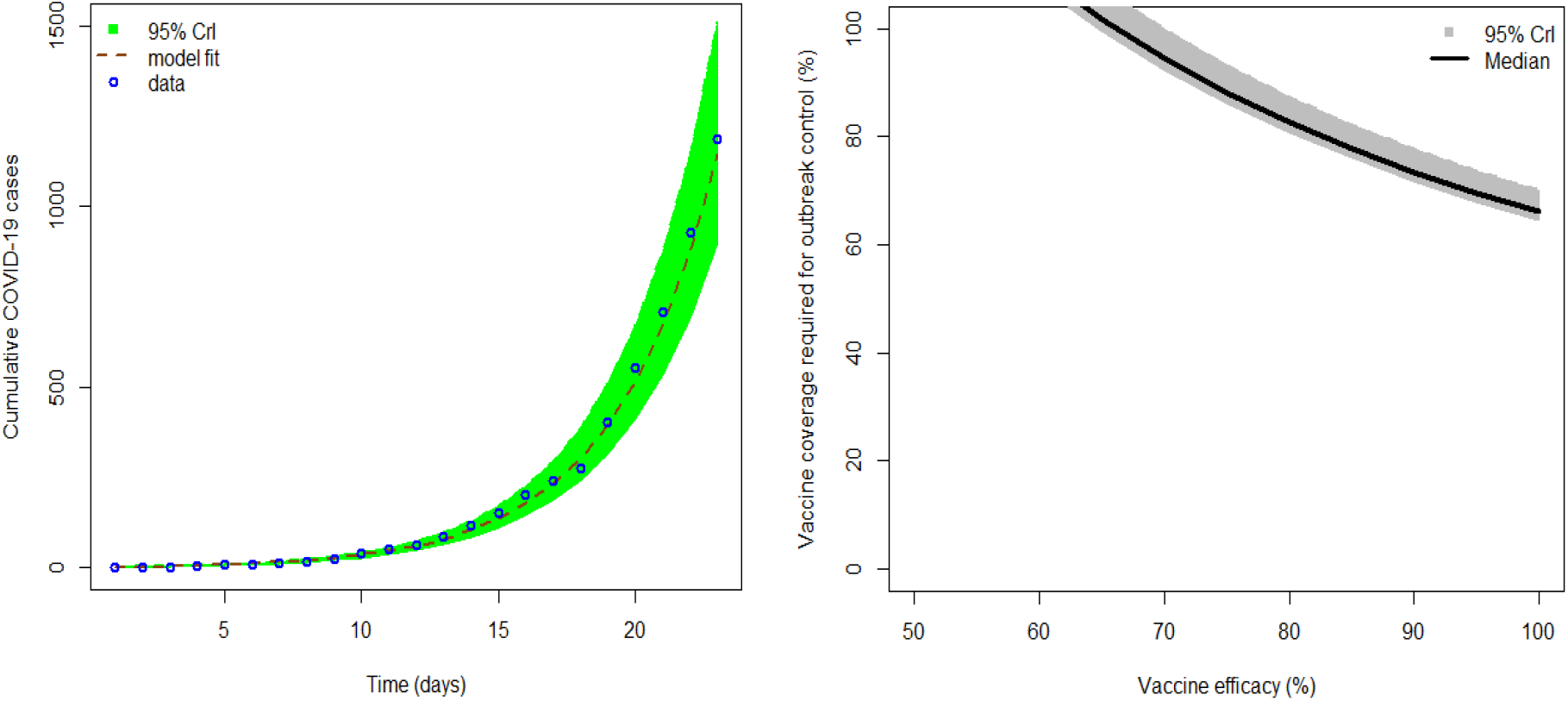
The first graph shows COVID-19 model fitting to cumulative cases where the green region is the 95% CrIs, the dashed brown line is the best model fit and the blue circles are the reported data for the cumulative number of COVID-19 cases in South Africa. The second graph shows the sensitivity analysis plot showing different vaccination coverages for different COVID-19 vaccine efficacy for South Africa. The dark grey regions are the 95% CrIs and the black line is the median.

With COVID-19 having been declared a global pandemic [9] and the urgent need to have an effective vaccine [10], there is a need to understand the utility of mass vaccination campaigns for this pandemic. Critical in the early stages of the disease is the need to clearly understand the spectrum of disease severity and transmission characteristics of the disease in order to identify optimal control measures. Many of the control measures suggested for this pandemic have been attributed to the lessons learnt in Wuhan, China [11]. The challenges associated with real-time analysis of an evolving epidemic are well articulated in [12]. These include testing capacity and delayed appearance of symptoms and asymptomatic carriage. The impact of COVID-19 on South Africa may differ from that on China and other regions such as Europe and North America. South Africa has unique circumstances, for example, it has the highest numbers of people living with HIV and one of the largest tuberculosis (TB) burdens in the world [13, 14]. Moreover, underlining disease conditions such as diabetes, hypertension and chronic obstructive pulmonary disease are prevalent in South African and these are known to present enormous challenges in the management of COVID-19 cases [15]. The age distribution for South Africa is also different from China and Europe as its young population accounts for the majority of the population [16].

Data from China and other settings have shown that SARS-CoV-2 is more infectious than influenza, and has a median incubation period of about 5 days and uncontrolled doubling time of 3 days [17, 18]. However, we have a limited understanding of the infectiousness of the virus in settings with different populations and a huge burden of other chronic conditions such as Africa. Mathematical models provide important insights in the understanding of emerging infectious diseases and informing public health policies. Several mathematical models have been used to understand COVID-19 transmission dynamics and inform public health policy [12, 19, 20, 21, 22]. The reproductive numbers of the COVID-19 epidemic in China have been determined in several modelling studies (Table 1).

**Table 1:** COVID-19 reproductive numbers from modelling studies in China.

| Study | Location | Reproductive number estimate |
| --- | --- | --- |
| Wu *et al* [28] | Wuhan | 2.68 [2.47-2.86] |
| Shen *et al.* [29] | Hubei province | 6.49 [6.31-6.66] |
| Liu *et al.* [30] | China and overseas | 2.90 [ 2.32-3.63] |
| Majumder and Mandl [31] | Wuhan | 2.55 [2.00-3.10] |
| Read *et al.* [32] | China | 3.11 [2.39-4.13] |
| Zhao *et al.* [33] | China | 2.24 [1.96-2.55] |
| Qun *et al.* [34] | China | 2.2 [1.40-3.90] |
| Tang *et al.* [35] | China | 6.47 [ 5.71-7.23] |
| Imai *et al.* [36] | Wuhan | 2.5 [1.50-3.50] |

Here, we adapt a susceptible-exposed-infected-removed (*SEIR*) compartmental model to quantify early transmissibility of COVID-19 in South Africa and explore the potential utility of a vaccine in containing the disease. The *SEIR* model has been used to model respiratory infections including Middle East Respiratory Syndrome (MERS) [23, 24], COVID-19 in Wuhan China [11], influenza [25, 26] and global tracking of COVID-19 [27]. In addition, we estimate the reduction in effective contacts after the implementation of the severe and extreme shutdown of the society, and this is critical in determining the impact of social distancing in the South African context.

## Methods

We use a standard deterministic compartmental *SEIR* model to simulate COVID-19 in South Africa. The model classifies the human population into four epidemiological compartments at any time *t*, the susceptible *S*(*t*), exposed *E*(*t*), infected *I*(*t*) and the recovered *R*(*t*). The total population is thus given by *N*(*t*) = *S*(*t*) + *E*(*t*) + *I*(*t*) + *R*(*t*). Susceptible individuals are infected upon interaction with infectious COVID-19 individuals and the rate of daily generation of newly infected cases is given by *λ*(*t*) = *βS*(*t*)*I*(*t*)/*N*, where the parameter *β* is the effective contact rate, *i.e*. the contact that will result in an infection. Exposed individuals in the *E*(*t*) compartment become infectious at a constant rate *σ* and move to the *I*(*t*) class. Infected individuals *I*(*t*), recover at a constant rate *γ* to the removed class *R*(*t*). The schematic model flow diagram is presented in *Supplementary Figure 1*. The model assumptions result in the following system of differential equations.

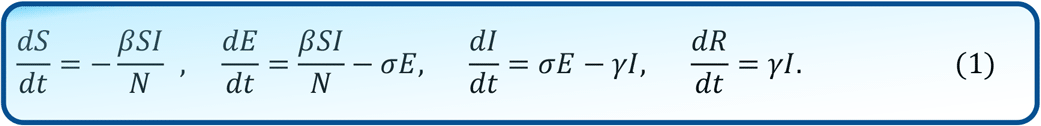

Following a similar approach in [37], we use a Markov Chain Monte Carlo (MCMC) within a Bayesian framework (in R FME package [38]) to fit the model to the cumulative data of confirmed COVID-19 cases in South Africa and estimate the magnitude of the epidemic using the basic reproductive number and quantify required vaccines’ attributes to stem similar outbreaks. We used data on COVID-19 cases published by the South African Department of Health from the 5^th^ of March 2020 to 28 March 2020 prior to the *lockdown* to estimate the basic reproductive number [39]. In addition, we estimate the percentage reduction in effective contacts after the *lockdown* ∈ by fitting the model to cumulative COVID-19 cases reported a week after the *lockdown* (from 29^th^ March to 11^th^ April 2020). Here ∈ ∈ (0,1) with ∈ ≃ 0 implying an ineffective *lockdown* while ∈ ≃ 1 implying a completely effective *lockdown*. The effective contact reduction term multiplies the effective contact rate in the model to give (1 − ∈)*β*.

In the fitting, we estimated *β, σ, γ* (within parameter ranges in *Supplementary Table 1*) and varied the initial infected population. Gaussian likelihood was used to draw model parameter posteriors assuming uniform non-informative priors while the variances were regarded as nuisance parameters. The MCMC chain was generated with at least 100000 runs for the final fitting excluding the burn-in period. Chain convergence was examined visually and using the Coda R package [40]. Uncertainty of each estimated parameter was evaluated by analysing the MCMC chains and calculating the 2.5 and 97.5 quantiles of the chain around its median to give the 95% credible interval (CrI).

The basic reproductive number (ℛ_0_), is as a measure of the average number of secondary cases generated by a primary case and is an important statistic for quantifying intervention programmes [41, 42]. Using an intuitive mathematical approach, the reproductive number of model system (1) is given by ℛ_0_ = β/γ. The corresponding minimum vaccination coverage (*c*) for COVID-19 vaccine for different vaccine efficacy scenarios was estimated using the mathematical expression 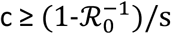 where *s* the proportional reduction of the susceptibility for individuals partially immunized.

## Results

Estimates of effective contact rate (*β*), the incubation period (1/*σ*), infectious period (1/*γ*), the percentage reduction in effective contacts (*ϵ*) and the basic reproductive number, ℛ_0_ for South Africa are shown in Table 2. The mathematical model (of the *SEIR* type) was fitted to the cumulative COVID-19 cases for South Africa at the national level (Figure 3). We estimated an effective contact rate 1.30 (95% Crl 1.21-1.39) per day, incubation period of 3.21 days (95% Crl 3.04-3.44 days), infectious period of 2.27 days (95% Crl 2.04-2.74 days) and ℛ_0_ of 2.95 (95% Crl 2.83-3.33) before the *lockdown*. The result ℛ_0_ >1 clearly shows disease sustainability in the country.

**Table 2:** Estimates of effective contact rate (*β*), the incubation period (1/*σ*), infectious period (1/*γ*), the percentage reduction in effective contacts (*ϵ*) and basic reproductive number (ℛ_0_).

| Parameter description | Estimated values [95 % CrIs] |
| --- | --- |
| **Before lockdown** |  |
| Effective contact rate ($\beta$) | 1.30 [1.21-1.39] day<sup>-1</sup> |
| Incubation period ($1/\sigma$) | 3.21 [3.04-3.44] days |
| Infectious period ($1/\gamma$) | 2.27 [2.04-2.74] days |
| Basic reproductive number ($\mathcal{R}_0$) | 2.95 [2.83-3.33] |
| **After lockdown** |  |
| Percentage reduction in effective contacts ($\epsilon$) | 80.31 [79.76-80.85]% |

**Figure 3:**
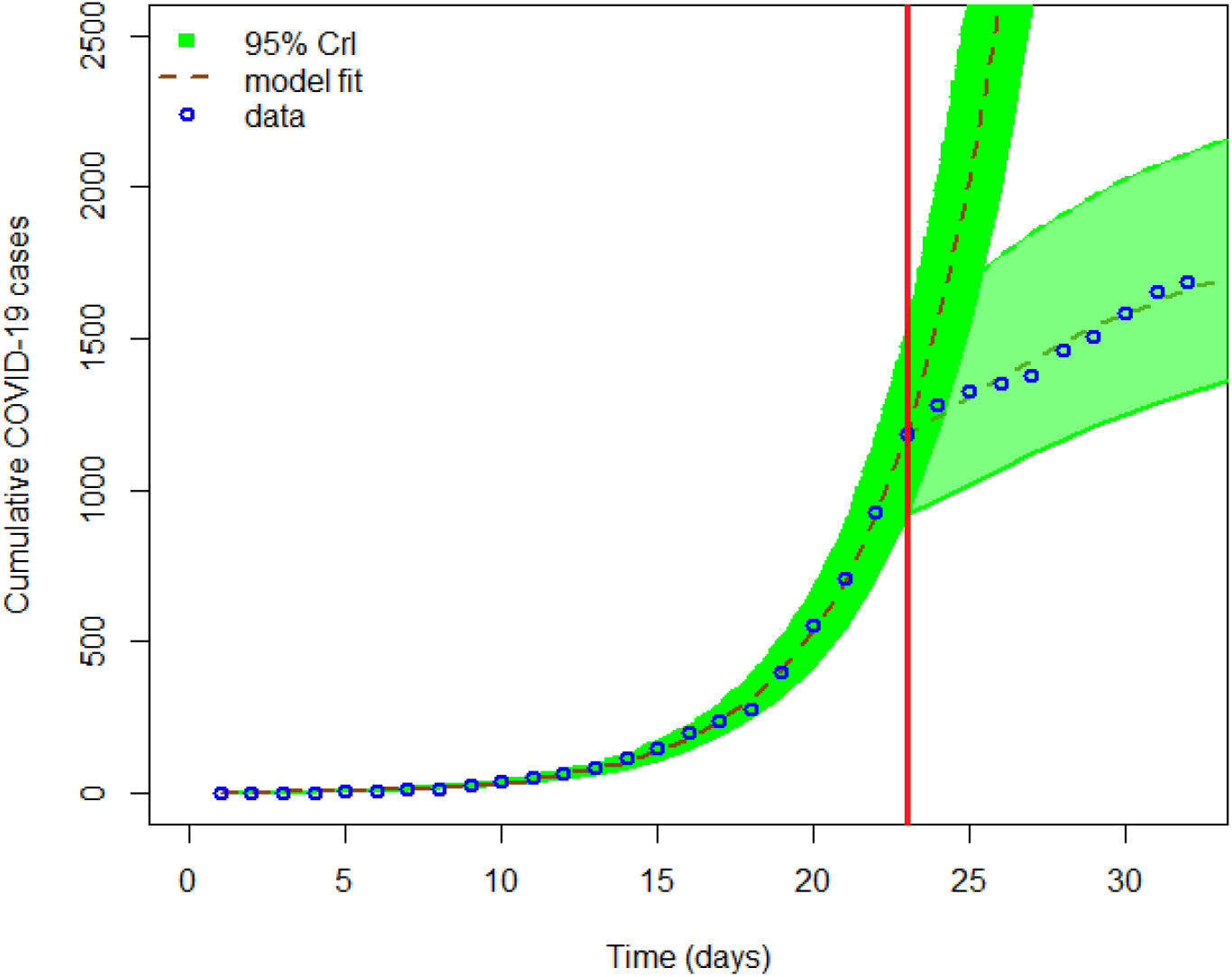
The first graph shows COVID-19 model fitting to cumulative cases where the green region is the 95% CrIs, the dashed brown line is the best model fit and the blue circles mark are the reported data for the cumulative number of COVID-19 cases in South Africa. The red vertical line denotes the time when the *lockdown* was implemented by the government. The dark green region show the trajectory the epidemic would have taken if a *lockdown* was not implemented and the light green region show the trajectory of the epidemic after the implementation of a *lockdown*.

Estimates of ℛ_0_ were used to conduct sensitivity analysis based on different COVID-19 vaccines’ efficacy assumptions to explore possible scenarios that may arise from mass vaccination campaigns, as scientists attempt to develop effective vaccines for COVID-19 [43, 44]. The vaccine efficacy scenarios were assumed to vary in the range of 50-100% (Figure 2). The results suggest that a vaccine with > 70% efficacy could have the potential to contain the COVID-19 outbreak in South Africa but at extremely high vaccination coverage rates of 94.44% (95% Crl 92.44-99.92%). As expected, vaccination coverage for epidemic control decreases with an increase in vaccine efficacy, with a vaccine of 100% efficacy requiring 66.10% (95% Crl 64.71-69.95%) coverage which is achievable under routine immunisation campaigns [45].

We also quantified the percentage reduction in effective contacts as a result of the *lockdown* mandated by the government of South Africa. Figure 3 shows that the epidemic is slowing down after the implementation of a *lockdown*. The results showed that the *lockdown* resulted in 80.31% (95% Crl 79.76-80.85%) reduction in effective contacts (Table 2) and consequently resulted in a reduction in the number of COVID-19 cases reported in the first two weeks of implementation. This confirms results in China that demonstrated the importance of quarantine, social distancing, and isolation in containing the pandemic [46].

## Discussion

COVID-19 has spread rapidly globally assisted by air travel in an increasingly connected world [47, 48]. Globally most countries, including South Africa, have adopted one form or another of the *lockdown* approaches in an attempt to curb disease transmission within their borders [11, 49, 50]. Our model estimate of ℛ_0_>1 confirms COVID-19 persistence in South Africa and indicate that the outbreak has the momentum to rapidly spread and spill over to other geographic regions of the country, in particular if the coming winter season (May to July) presents ideal environmental conditions for persistence of the virus. The estimate of ℛ_0_ for South Africa is in a similar range published for COVID-19 in other modelling studies (Table 1).

Hypothetical scenarios on vaccine efficacy demonstrated that, a vaccine of at least 70% efficacy would have been sufficient to contain the spread of COVID-19 in South Africa although at high vaccination coverage. However, it is important to note that expectations that the development of a highly effective vaccine for the novel-coronavirus will be achieved in the coming months are extremely optimistic, especially when considering that a vaccine has still not been successfully created for viruses like HIV, which has been in development for many years [44, 51]. Nevertheless, the huge global interest in quickly identifying an effective vaccine could increase the possibility that a successful vaccine candidate can be developed in the coming months [43]. Even when a safe and effective vaccine becomes available, there are several logistical and operational challenges that need to be addressed for successful deployment and for the vaccine to achieve the desired coverage [52, 53].

The modelled *lockdown* demonstrated 80.31% reduction in effective contacts, showing that it is an effective measure to bring the disease under control. However, the reduction in the number of daily reported cases should be interpreted with caution as this could also have been a result of many other factors such as reduced international travel to high-risk regions and behaviour change. As the epidemic continues to unfold, it remains to be seen what trend the epidemic will follow if local transmissions are sustained within South Africa. The implementation of this society shut down is not sustainable in the long term, nor is likely to be tolerated for too long by the population. A vaccine would be an ideal preventative strategy for COVID-19 but it appears that it should be complemented with prevention approaches such as isolation, quarantine, personal hygiene and limitations of public gatherings in order to achieve optimal protection of the population in South Africa.

The economic and social burden of the disease continues to be felt and this likely to be enhanced by an extension of the current *lockdown* of 21 days by a further 2 weeks [54]. However it is unclear whether a *stringent lockdown* could be maintained for a longer period given the socio-economic challenges of the country where a significant percent of adults are involved in informal employment and others have jobs that do not allow them to work from home. This could affect the effectiveness of the *lockdown* in many of the townships as individuals will have to relax *lockdown* conditions in order to be economically active and prevent financial woes on individuals in urban communities. While the epidemic seems to have slowed down as a results of the *lockdown* (Figure 3), there is need for continued scientific investigation including explorations through mathematical models to monitor the trend with the aim of informing public health policy in the short-term.

The study has some limitations. The estimate of ℛ_0_ is based on available data and this estimate could possibly change depending on the quality of the data from the start of the epidemic (with possible under-reporting of cases in the initial phases of the epidemic). We note that spatial modelling mainly in the affected provinces would have been ideal but we did not have good data on a finer resolution to effectively parameterise a spatial model but as the epidemic evolves, nascent data on local COVID-19 transmission in South Africa is becoming available. We used a simple mathematical model without other population demographics as these were not important for short-term prediction [55] and such models are also important when epidemiological and clinical disease characteristics of the disease are not well established as is the case for COVID-19 [56]. The simple model is only intended to give preliminary estimates for an epidemic that is evolving and whose trend has the potential to change dramatically overtime. However, it would be interesting to see how our results will change when a more complicated model is used. Despite these shortfalls, findings in this study are important in understanding the transmissibility of the virus and informing the development of COVID-19 prevention and control programmes in South Africa and outlining mass vaccination expectations.

COVID-19 pandemic has continued to spread and causing many deaths than any infectious disease we have seen in recent years and this calls for an urgent and well-coordinated timely and effective public health response. Currently there is no proven treatment or vaccines for COVID-19 and countries have embraced quarantine, social distancing, and isolation of infected individuals as to contain the pandemic. Thus, the building of suitable mathematical models to weight out the impact of current public health control measures and explore the potential utility of anticipated biomedical interventions such as vaccines is paramount.

## Data Availability

The manuscript include all data required.

## Author Contributions

ZM, FN, NM and GM conducted model analysis and were responsible initial manuscript preparation. DFC, TS assisted with the conceptualization of the project and preparation of the manuscript. All authors were involved in writing the manuscript.

## Ethical Approval

Ethics approval was not required for this study.

## Supplementary Material

**Supplementary Figure 1:**
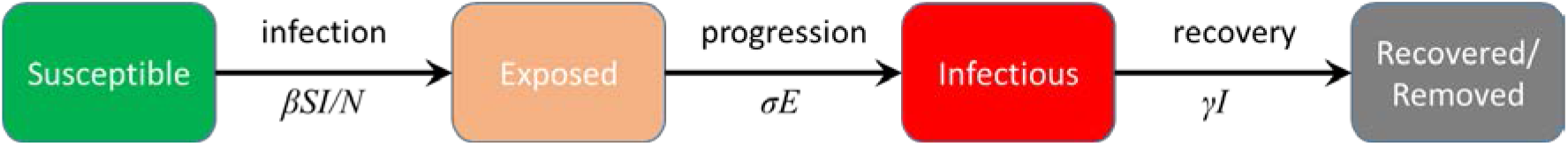
Schematic COVID-19 model diagram outlining infection progression. The arrows connecting compartments denote COVID-19 infection at rate *βS*(*t*)*I*(*t*)/*N*, progression to infectiousness *σE* and recovery rate *γI* respectively.

**Supplementary Table 1:**
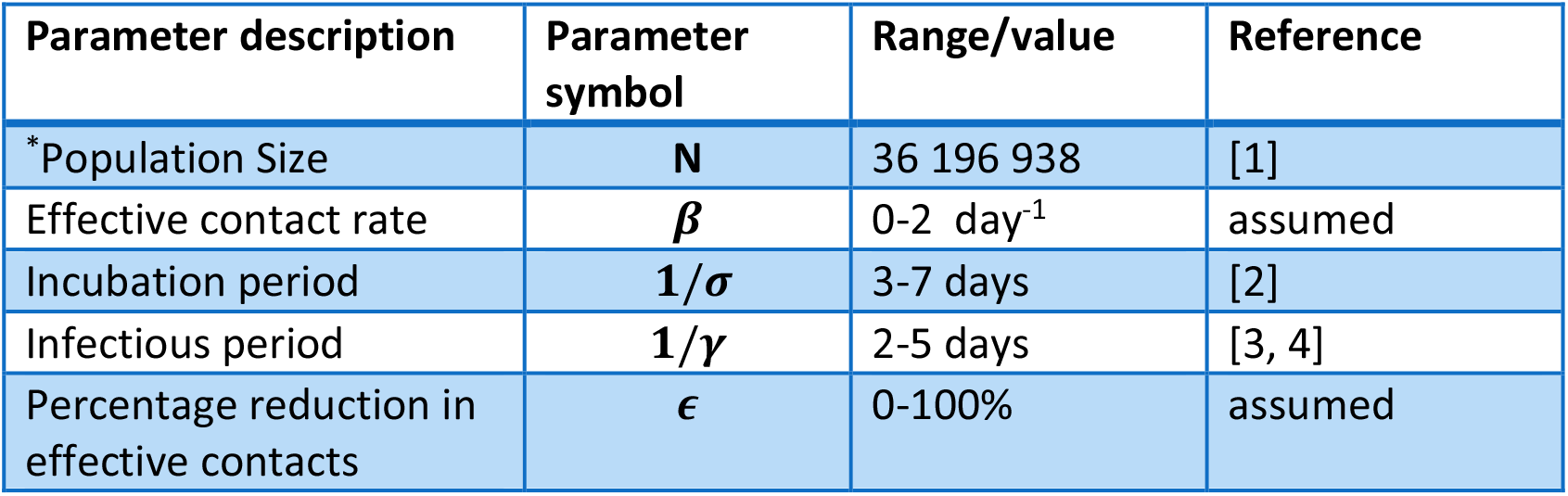
COVID-19 model parameters and values. *We used population size for Gauteng, Western Cape, KwaZulu-Natal and Free State provinces in estimating national level estimates as these were the only provinces with COVID-19 cases in the early phases of the epidemic. Population estimates were based on the 2019 census [1].

| Parameter description | Parameter symbol | Range/value | Reference |
| --- | --- | --- | --- |
| *Population Size | **N** | 36 196 938 | [1] |
| Effective contact rate | **$\beta$** | 0-2 day<sup>-1</sup> | assumed |
| Incubation period | **$1/\sigma$** | 3-7 days | [2] |
| Infectious period | **$1/\gamma$** | 2-5 days | [3, 4] |
| Percentage reduction in effective contacts | **$\epsilon$** | 0-100% | assumed |

